# No Evidence that Ongoing HIV-Specific Immune Responses Contribute to Persistent Inflammation and Immune Activation in Persons on Long-Term ART

**DOI:** 10.1101/2021.11.08.21266096

**Authors:** Adam R. Ward, Allison S. Thomas, Eva M. Stevenson, Szu-Han Huang, Sheila M. Keating, Rajesh T. Gandhi, Deborah K. McMahon, Ronald J. Bosch, Bernard J. Macatangay, Joshua C. Cyktor, Joseph J. Eron, John W. Mellors, R. Brad Jones, for the ACTG A5321 Team

## Abstract

**Background:** People with HIV (PWH) have persistently elevated levels of inflammation and immune activation despite suppressive antiretroviral therapy (ART), with specific biomarkers showing associations with non-AIDS-defining morbidities and mortality. We investigated the potential role of the HIV-specific adaptive immune response, which also persists under ART, in driving levels of these clinically relevant biomarkers.

**Methods:** HIV-specific T-cell responses and antibody concentrations were measured at study entry in the ACTG A5321 cohort, following a median of 7 years of suppressive ART. HIV persistence measures including cell-associated (CA)-DNA, CA-RNA, and plasma HIV RNA (single-copy assay) were also assessed. Plasma inflammatory biomarkers and T-cell activation and cycling were measured at a pre-ART time point and at study entry.

**Results:** Neither the magnitudes of HIV-specific T-cell responses nor HIV antibody levels were correlated with levels of the inflammatory or immune activation biomarkers, including hs-CRP, IL-6, neopterin, sCD14, sCD163, %CD38^+^HLA-DR^+^ CD8^+^ and CD4^+^ cells, and %Ki67^+^ CD8^+^ and CD4^+^ cells – including after adjustment for pre-ART biomarker level. Magnitudes of T-cell responses to HIV-Pol were correlated with TNF-α levels, but this was confounded by several factors. Plasma HIV RNA levels were correlated with CD8^+^ T-cell activation (r = 0.25, p = 0.027), but other HIV persistence parameters were not associated with these biomarkers. In mediation analysis, relationships between HIV persistence parameters and inflammatory biomarkers were not influenced by either HIV-specific T-cell responses or antibody levels.

**Conclusions:** Adaptive HIV-specific immune responses do not appear to contribute to the elevated inflammatory and immune activation profile associated with morbidity and mortality under long-term ART.

**Summary:** HIV-specific T-cell and antibody responses persist over years of suppressive ART, but there is no evidence that these ongoing immune responses contribute to elevated levels of inflammation and immune activation in people living with HIV on long-term ART.

## Introduction

Antiretroviral therapy (ART) can durably suppress human immunodeficiency virus (HIV) viremia in people with HIV (PWH), yet despite effective ART and host antiviral immune responses HIV persists in a reservoir of infected cells necessitating lifelong treatment. ART reduces systemic inflammation and immune activation, resulting in a tremendous decline in AIDS-related morbidity and mortality. Yet, even in the setting of long-term, well-controlled HIV infection, PWH on ART demonstrate elevated levels of inflammation and immune activation – especially when ART was initiated in chronic infection – which predict a broad array of morbidities and increased mortality [1–5].

The etiology of persistent inflammation and immune activation under ART is incompletely understood, though multiple potential mechanisms likely contribute, including increased intestinal permeability and microbial translocation [6–8], and co-infections such as cytomegalovirus (CMV) [9,10]. A role has been proposed for HIV reservoirs [11], though we have previously shown that markers of HIV persistence are not associated with inflammation or immune activation during long-term ART [12,13]. We have also previously shown that HIV-specific T-cell and antibody responses persist under long-term ART and are associated with HIV DNA levels [14–16]. However, it is not known if these on-ART HIV-specific immune responses play a role in driving persistent inflammation and immune activation.

Here, we sought to determine wither ongoing HIV-specific immune responses contribute to levels of clinically relevant inflammatory and immune activation (T-cell activation and cycling) biomarkers – selected for analysis based on prior reports of associations with morbidities and/or mortality [1–5] – in PWH on long-term ART.

## Methods

### Study Design and Approval

Data for this manuscript were collected from a longitudinal cohort of participants who initiated ART during viremic chronic HIV infection in AIDS Clinical Trials Group (ACTG) trials for treatment-naive individuals and enrolled in the ACTG HIV Reservoirs Cohort Study (A5321) [12]. Participants had no reported ART interruptions, with plasma HIV RNA levels <50 copies/mL by commercial assays at or before week 48 of ART and at all subsequent time points (isolated measurements <200 copies/mL were allowed). Clinical data were available from pre-ART and on-ART study visits. The institutional review boards at the authors’ institutions approved the study, and informed written consent was obtained from all participants.

### Virologic Assays

HIV cell-associated (CA) DNA (CA-DNA) and RNA (CA-RNA) were measured by quantitative PCR (qPCR) assays in peripheral blood mononuclear cells (PBMCs) using previously described methods [17]. CA-DNA and CA-RNA values per million CD4^+^ T-cells were calculated by dividing the total CA-DNA or CA-RNA copies/million PBMCs (normalized for CCR5 copies measured by qPCR [17]) by the CD4^+^ T-cell percentage (× 0.01) reported from the same specimen date or from a CD4^+^ T-cell percentage imputed using linear interpolation from specimen dates before and after the CA-DNA or CA-RNA results. Cell-free HIV RNA was quantified by integrase single-copy assay (iSCA) in blood plasma (5 mL) [18].

### Immunologic Assays

Levels of soluble biomarkers were evaluated from frozen plasma samples (analyzed in batch for each participant) as previously described [12]. In brief, plasma concentrations of high-sensitivity C-reactive protein (hs-CRP), interleukin 6 (IL-6), interferon-gamma-induced protein 10 (IP-10), neopterin, soluble CD14 (sCD14), soluble CD163 (sCD163), and tumor necrosis factor alpha (TNF-α) were quantified using enzyme-linked immunosorbent assay (ELISA) kits per manufacturer’s instructions (R&D, Minneapolis, MN). Levels of T-cell activation and cell-cycling biomarkers in cryopreserved PBMCs from each participant were determined in batch using multicolor flow cytometry.

### IFN-γ ELISPOT Assays

Interferon-gamma (IFN-γ) enzyme-linked immune absorbent spot (ELISPOT) assays against HIV-gene product peptide pools and a CMV-pp65 peptide pool were performed as previously described [14]. In brief, Multiscreen IP 96-well plates (Millipore) were coated with 0.5 μg/mL of anti-IFN-γ antibody (clone 1-D1K, Mabtech, Sweden) in phosphate-buffered saline and incubated overnight. Plates were washed, PBMCs were added at 2 × 10^5^ cells per well, and HIV peptide pools or CMV-pp65 peptide pool (10 μg/mL/peptide) and phytohemagglutinin (2 μg/mL) were added. Plates were incubated overnight, washed and secondary antibody was added (clone 7-B6-1, Mabtech) and incubated for 1 hour. Plates were developed with Streptavidin-ALP (Mabtech) and with Color Development Buffer (Bio-Rad, Hercules, CA). Plates were washed, dried overnight and spots were counted.

### HIV Antibody Assays

HIV antibodies were measured as previously described [15]. In brief, Less-sensitive and Avidity-modified VITROS HIV 1 + 2 were used to measure antibodies against HIV envelope (Env) [19].

### Statistics

Statistical analyses including univariate statistics and nonparametric Spearman correlations and partial correlations (adjusting for other variables) were performed in SAS v.9.4 (SAS Institute Inc., Cary, NC). Structural equation modeling (SEM) for mediation analysis was performed using the SAS CALIS procedure, with reported coefficients representing standardized effects; variables in SEM models were rank-transformed [20] using the SAS RANK procedure to match the nonparametric Spearman correlation approach. For hs-CRP values above the limit of assay detection (10,000 ng/mL), values were imputed as 10,001 ng/mL and analyzed as the highest rank. For CA-RNA values below the limit of assay detection (13.6 copies/million CD4^+^ T-cells), values were imputed as 13 copies/million CD4^+^ T-cells and analyzed as the lowest rank. For plasma HIV RNA via iSCA values below the limit of assay detection (0.7 copies/mL), values were imputed as 0.5 copies/mL and analyzed as the lowest rank.

## Results

### Study Population

We previously assessed HIV-specific T-cell responses, along with CMV-pp65-specific T-cell responses, by IFN-γ ELISPOT in 99 participants from the ACTG A5321 cohort [14]. Participants initiated ART during chronic HIV infection and had subsequent well-documented, sustained virologic suppression prior to study entry (**Figure S1**) and throughout the study period [12,16]. Responses were measured at A5321 study entry, a median of 7 (range 4 to 15) years after ART initiation, by IFN-γ ELISPOT assays with peptide pools spanning: i) HIV-Gag, ii) HIV-Env, iii) HIV-Pol, iv) HIV-Nef/Tat/Rev (combined peptide pool), and v) CMV-pp65. In this previous study as well as in a follow-up longitudinal study, ELISPOT responses were background subtracted (thus, nonzero responses were >1x background), but no other ad hoc empirical cutoff was applied [14,16]. We also previously measured antibody levels against HIV-Env in 101 participants from the A5321 cohort at study entry [15], with an overlap of 80 participants between those with T-cell response measurements and HIV antibody level measurements. Participant characteristics relevant to the current study are provided in **Tables 1 and S1**.

**Table 1.**
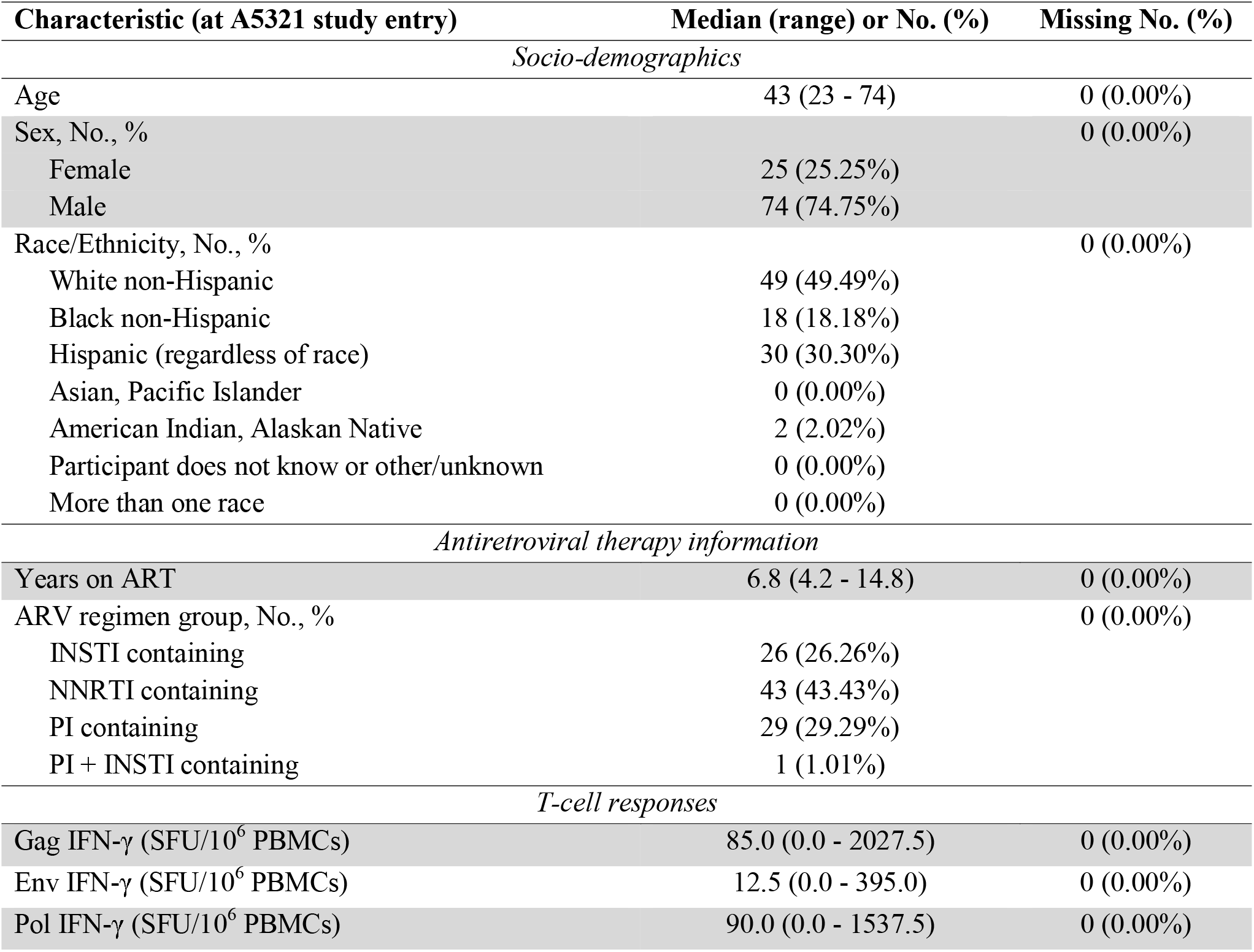
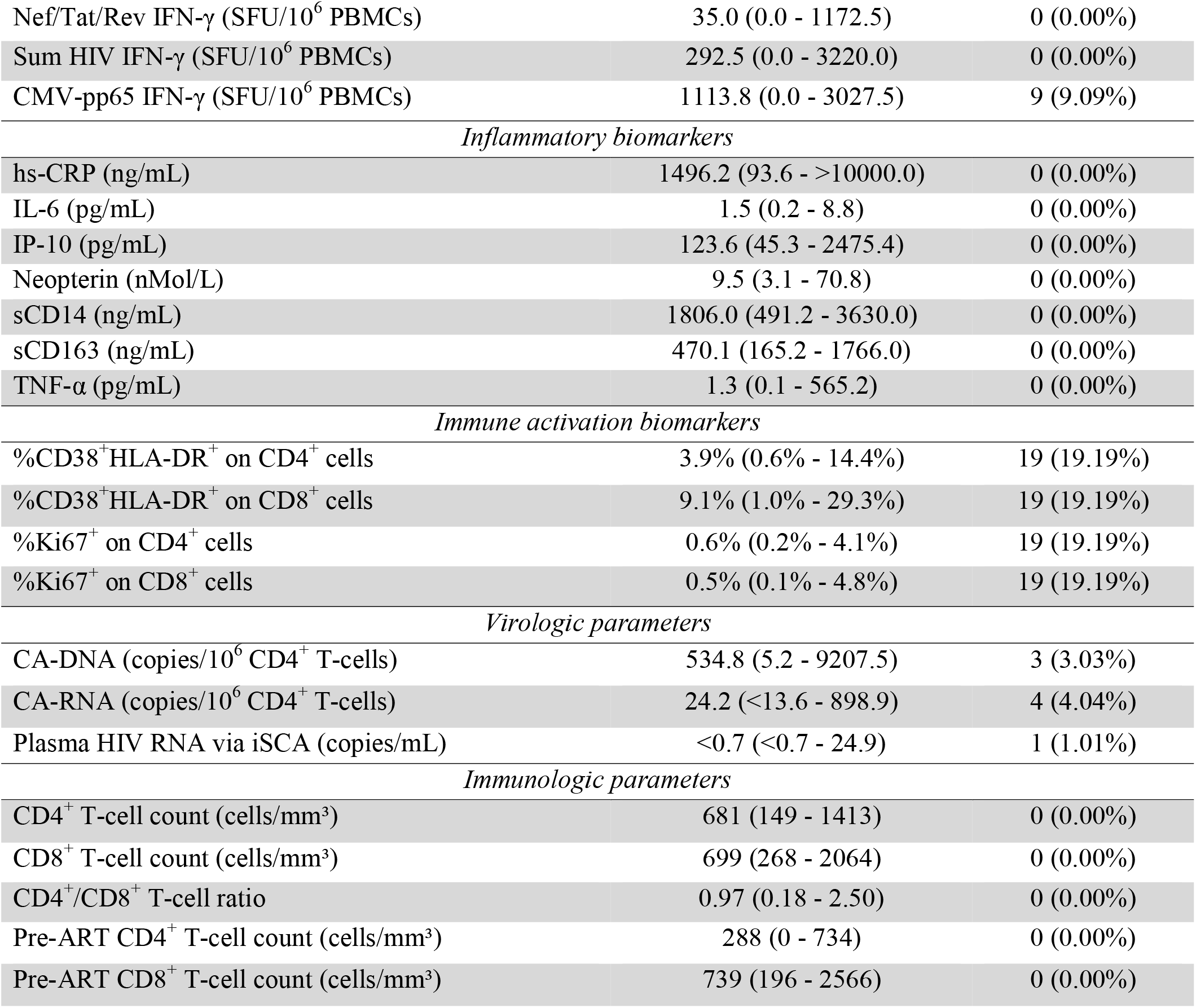
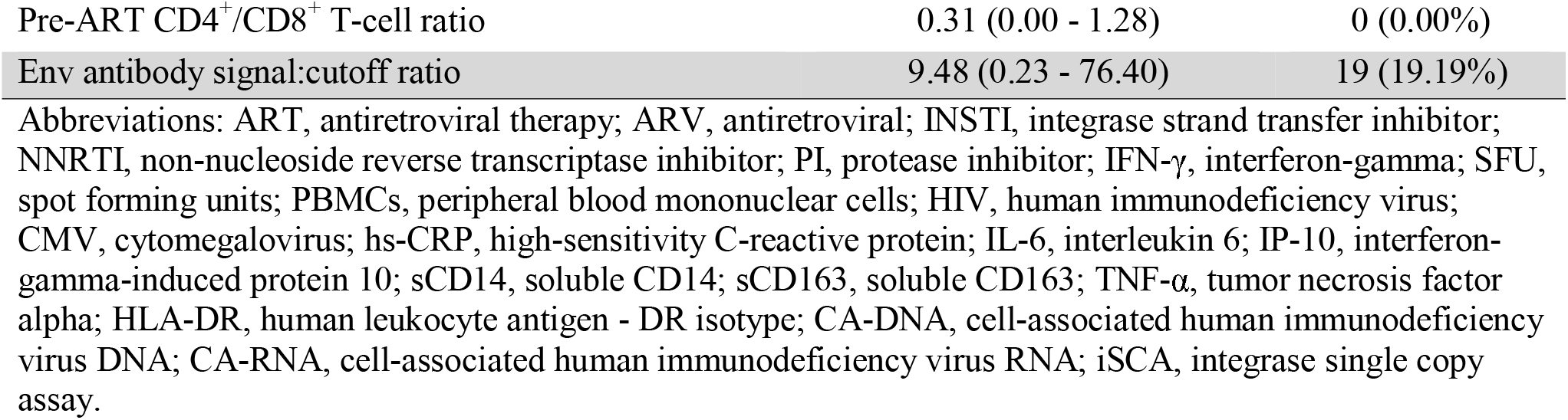
Participant characteristics (n=99)

| Characteristic (at A5321 study entry) | Median (range) or No. (%) | Missing No. (%) |
| --- | --- | --- |
| *Socio-demographics* |  |  |
| Age | 43 (23 - 74) | 0 (0.00%) |
| Sex, No., % |  | 0 (0.00%) |
| Female | 25 (25.25%) |  |
| Male | 74 (74.75%) |  |
| Race/Ethnicity, No., % |  | 0 (0.00%) |
| White non-Hispanic | 49 (49.49%) |  |
| Black non-Hispanic | 18 (18.18%) |  |
| Hispanic (regardless of race) | 30 (30.30%) |  |
| Asian, Pacific Islander | 0 (0.00%) |  |
| American Indian, Alaskan Native | 2 (2.02%) |  |
| Participant does not know or other/unknown | 0 (0.00%) |  |
| More than one race | 0 (0.00%) |  |
| *Antiretroviral therapy information* |  |  |
| Years on ART | 6.8 (4.2 - 14.8) | 0 (0.00%) |
| ARV regimen group, No., % |  | 0 (0.00%) |
| INSTI containing | 26 (26.26%) |  |
| NNRTI containing | 43 (43.43%) |  |
| PI containing | 29 (29.29%) |  |
| PI + INSTI containing | 1 (1.01%) |  |
| *T-cell responses* |  |  |
| Gag IFN-γ (SFU/10<sup>6</sup> PBMCs) | 85.0 (0.0 - 2027.5) | 0 (0.00%) |
| Env IFN-γ (SFU/10<sup>6</sup> PBMCs) | 12.5 (0.0 - 395.0) | 0 (0.00%) |
| Pol IFN-γ (SFU/10<sup>6</sup> PBMCs) | 90.0 (0.0 - 1537.5) | 0 (0.00%) |
| Nef/Tat/Rev IFN-γ (SFU/10<sup>6</sup> PBMCs) | 35.0 (0.0 - 1172.5) | 0 (0.00%) |
| Sum HIV IFN-γ (SFU/10<sup>6</sup> PBMCs) | 292.5 (0.0 - 3220.0) | 0 (0.00%) |
| CMV-pp65 IFN-γ (SFU/10<sup>6</sup> PBMCs) | 1113.8 (0.0 - 3027.5) | 9 (9.09%) |
| *Inflammatory biomarkers* |  |  |
| hs-CRP (ng/mL) | 1496.2 (93.6 - >10000.0) | 0 (0.00%) |
| IL-6 (pg/mL) | 1.5 (0.2 - 8.8) | 0 (0.00%) |
| IP-10 (pg/mL) | 123.6 (45.3 - 2475.4) | 0 (0.00%) |
| Neopterin (nMol/L) | 9.5 (3.1 - 70.8) | 0 (0.00%) |
| sCD14 (ng/mL) | 1806.0 (491.2 - 3630.0) | 0 (0.00%) |
| sCD163 (ng/mL) | 470.1 (165.2 - 1766.0) | 0 (0.00%) |
| TNF-α (pg/mL) | 1.3 (0.1 - 565.2) | 0 (0.00%) |
| *Immune activation biomarkers* |  |  |
| %CD38<sup>+</sup>HLA-DR<sup>+</sup> on CD4<sup>+</sup> cells | 3.9% (0.6% - 14.4%) | 19 (19.19%) |
| %CD38<sup>+</sup>HLA-DR<sup>+</sup> on CD8<sup>+</sup> cells | 9.1% (1.0% - 29.3%) | 19 (19.19%) |
| %Ki67<sup>+</sup> on CD4<sup>+</sup> cells | 0.6% (0.2% - 4.1%) | 19 (19.19%) |
| %Ki67<sup>+</sup> on CD8<sup>+</sup> cells | 0.5% (0.1% - 4.8%) | 19 (19.19%) |
| *Virologic parameters* |  |  |
| CA-DNA (copies/10<sup>6</sup> CD4<sup>+</sup> T-cells) | 534.8 (5.2 - 9207.5) | 3 (3.03%) |
| CA-RNA (copies/10<sup>6</sup> CD4<sup>+</sup> T-cells) | 24.2 (<13.6 - 898.9) | 4 (4.04%) |
| Plasma HIV RNA via iSCA (copies/mL) | <0.7 (<0.7 - 24.9) | 1 (1.01%) |
| *Immunologic parameters* |  |  |
| CD4<sup>+</sup> T-cell count (cells/mm<sup>3</sup>) | 681 (149 - 1413) | 0 (0.00%) |
| CD8<sup>+</sup> T-cell count (cells/mm<sup>3</sup>) | 699 (268 - 2064) | 0 (0.00%) |
| CD4<sup>+</sup>/CD8<sup>+</sup> T-cell ratio | 0.97 (0.18 - 2.50) | 0 (0.00%) |
| Pre-ART CD4<sup>+</sup> T-cell count (cells/mm<sup>3</sup>) | 288 (0 - 734) | 0 (0.00%) |
| Pre-ART CD8<sup>+</sup> T-cell count (cells/mm<sup>3</sup>) | 739 (196 - 2566) | 0 (0.00%) |
| Pre-ART CD4<sup>+</sup>/CD8<sup>+</sup> T-cell ratio | 0.31 (0.00 - 1.28) | 0 (0.00%) |
| Env antibody signal:cutoff ratio | 9.48 (0.23 - 76.40) | 19 (19.19%) |

Abbreviations: ART, antiretroviral therapy; ARV, antiretroviral; INSTI, integrase strand transfer inhibitor; NNRTI, non-nucleoside reverse transcriptase inhibitor; PI, protease inhibitor; IFN-γ , interferon-gamma; SFU, spot forming units; PBMCs, peripheral blood mononuclear cells; HIV, human immunodeficiency virus; CMV, cytomegalovirus; hs-CRP, high-sensitivity C-reactive protein; IL-6, interleukin 6; IP-10, interferon-gamma-induced protein 10; sCD14, soluble CD14; sCD163, soluble CD163; TNF-α , tumor necrosis factor alpha; HLA-DR, human leukocyte antigen - DR isotype; CA-DNA, cell-associated human immunodeficiency virus DNA; CA-RNA, cell-associated human immunodeficiency virus RNA; iSCA, integrase single copy assay.

### Magnitudes of HIV-Specific T-cell Responses are not Cross-Sectionally Associated with Levels of Inflammation or Immune Activation

To determine whether IFN-γ-producing HIV-specific T-cell responses could be contributing to levels of inflammation and immune activation on-ART, we examined associations between magnitudes of these responses (measured at A5321 study entry) with levels of plasma inflammatory biomarkers (also measured at A5321 study entry), including hs-CRP, IL-6, IP-10, neopterin, sCD14, sCD163, and TNF-α, as well as with levels of immune activation biomarkers (measured at A5321 study entry), including %CD38^+^HLA-DR^+^ CD4^+^ cells, %CD38^+^HLA-DR^+^ CD8^+^ cells, %Ki67^+^ CD4^+^ cells, and %Ki67^+^ CD8^+^ cells. A major strength of this longitudinal cohort study is the measurement of pre-therapy levels of inflammatory and immune activation biomarkers, allowing us to adjust for pre-ART biomarker level in the above associations. By adjusting for the pre-ART biomarker level, we were able to determine if on-ART HIV-specific T-cell responses influence inflammation and immune activation regardless of a participant’s baseline level of the respective biomarker prior to ART initiation. Additionally, we controlled for potential confounders in the above associations as detailed below.

For on-ART inflammatory biomarkers, TNF-α levels were negatively associated with magnitudes of responses to HIV-Pol (r = −0.21, p = 0.040), however this association was weak and did not remain significant after controlling for potential joint confounding by pre-ART plasma viral load, CD4^+^ T-cell count, and CD4^+^/CD8^+^ T-cell ratio, years on ART at A5321 entry, and age at A5321 entry - suggesting that host/viral interactions prior to ART initiation led to the observed on-ART association. TNF-α levels were not associated with magnitudes of T-cell responses to any other HIV gene product or to CMV-pp65 (**Table 2**). No other inflammatory biomarker was associated with magnitudes of HIV-specific, or CMV-pp65-specific, T-cell responses, even after adjusting for pre-ART biomarker level (**Table 2**), nor were any of the measured immune activation biomarkers (**Table 2**). These results indicate that HIV-specific T-cell responses, as assessed by *ex vivo* IFN-γ production, do not appreciably contribute to on-ART inflammation and immune activation.

**Table 2.**
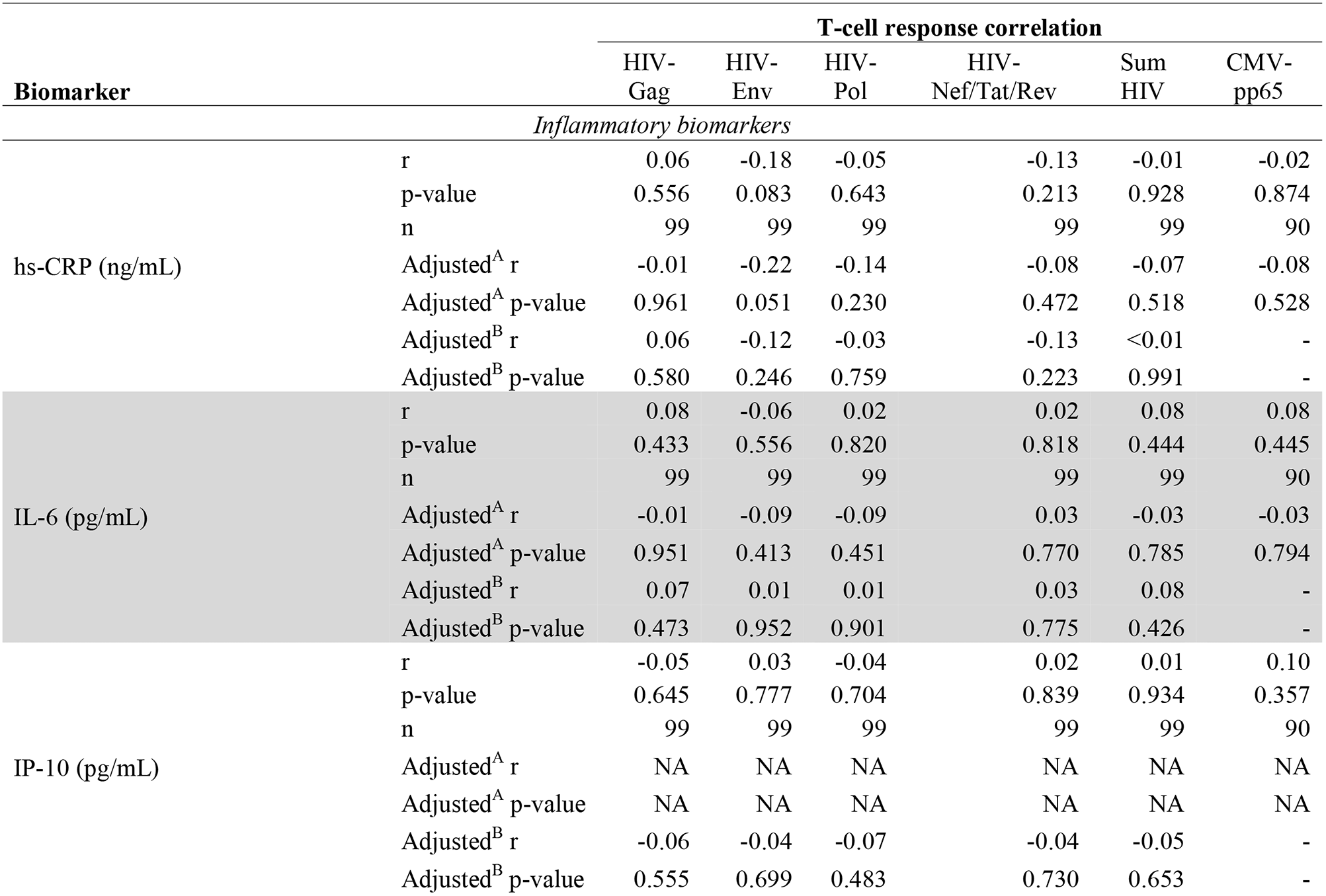
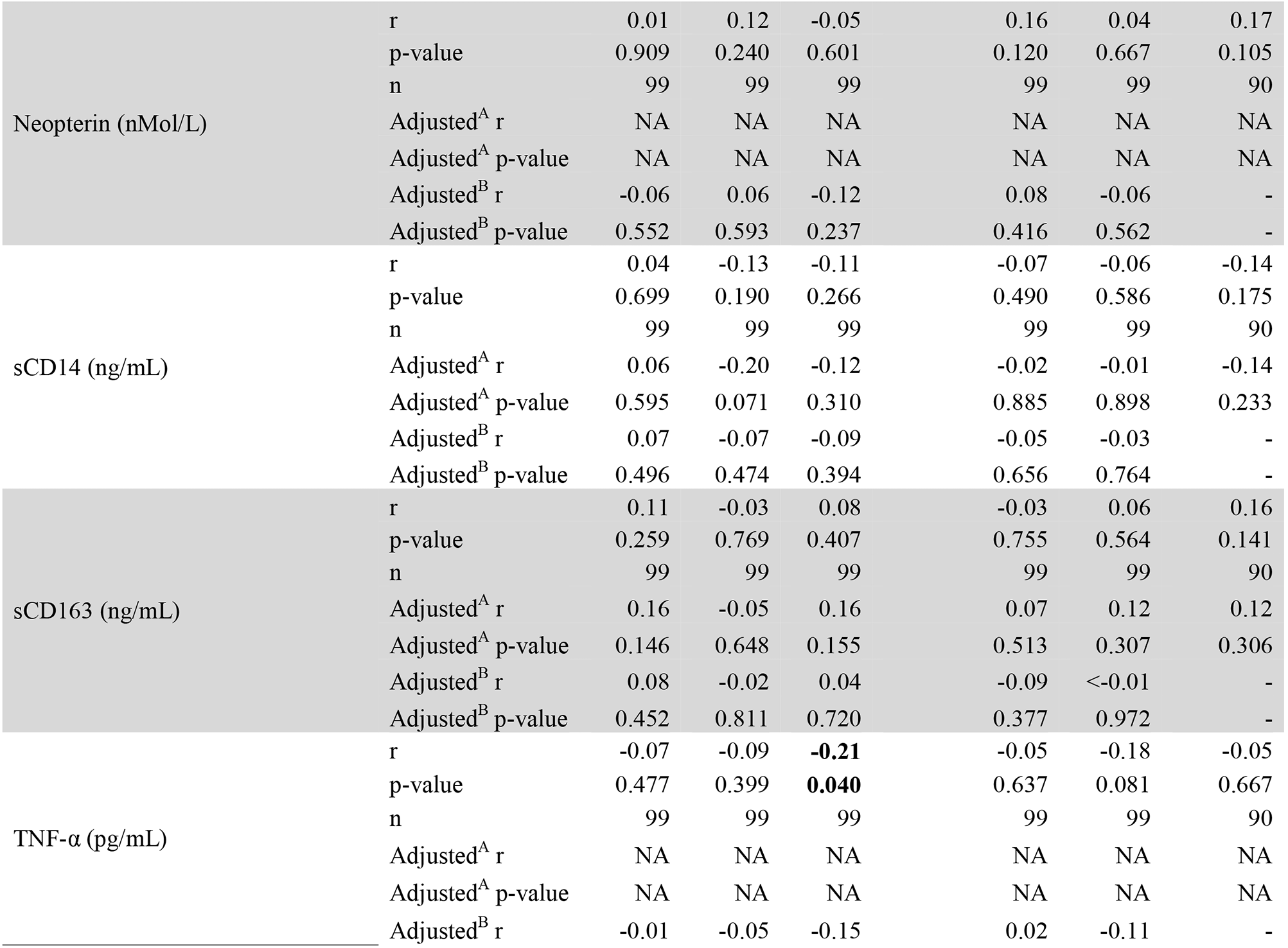
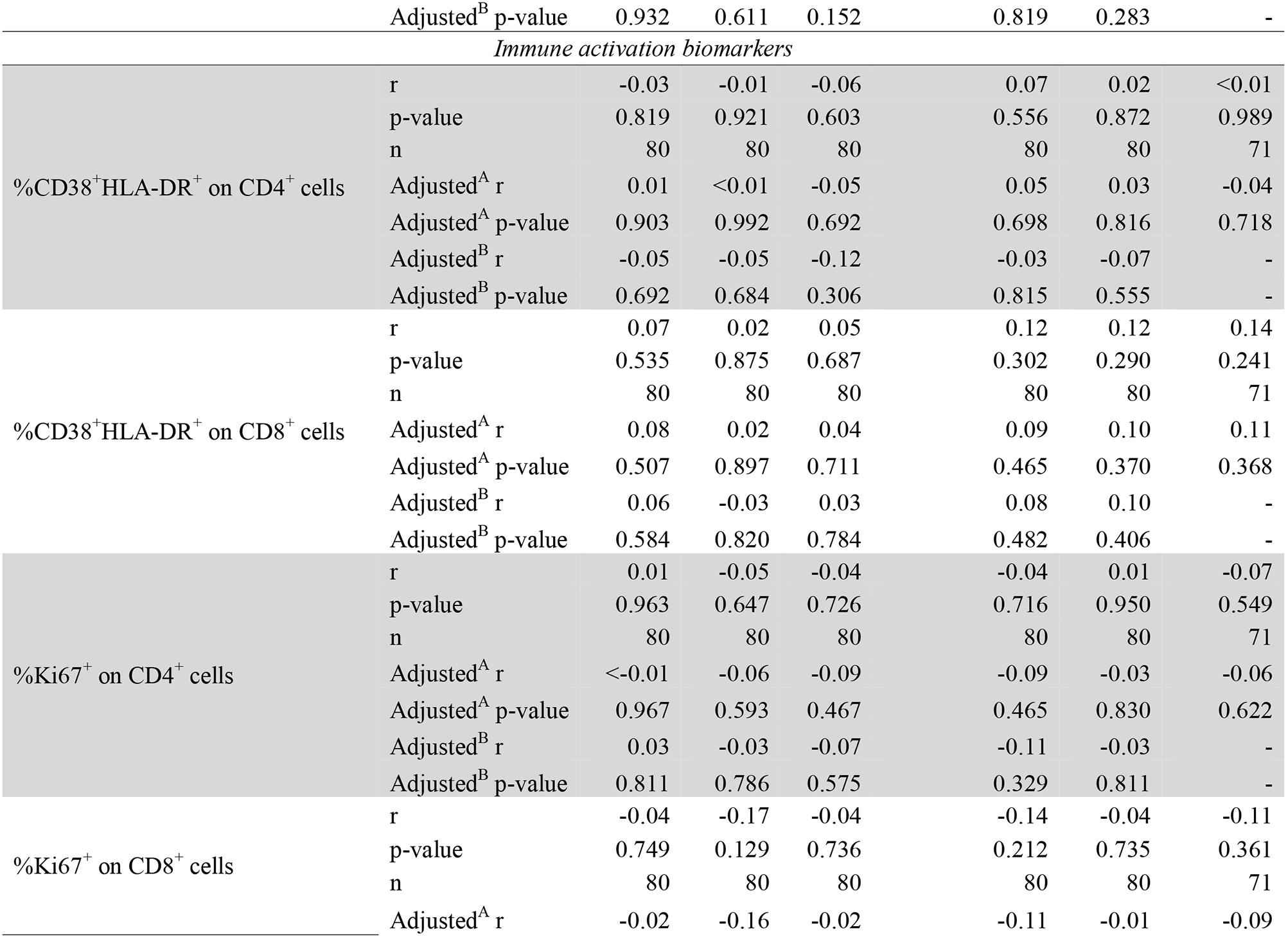
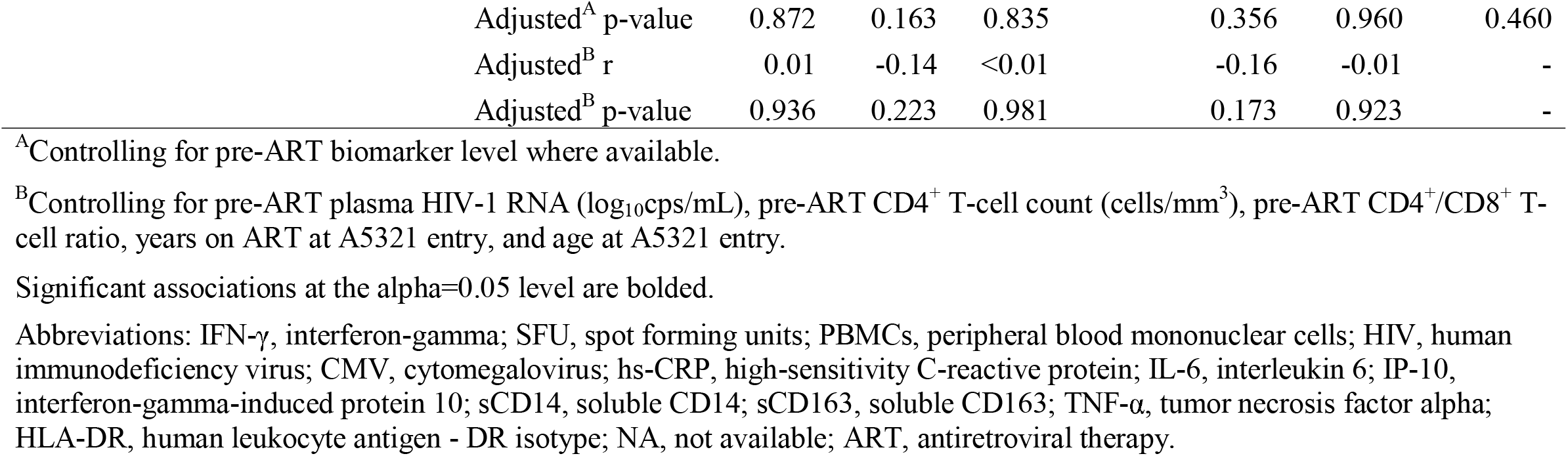
Spearman correlations between magnitudes of IFN-γ T-cell responses with inflammatory and immune activation biomarkers at A5321 study entry

### HIV Antibody Levels are not Associated with Levels of Inflammation or Immune Activation

We next asked if HIV antibody levels, another component of the ongoing adaptive immune response to HIV in treated infection [15], were associated with levels of inflammatory or immune activation biomarkers at A5321 study entry. No inflammatory biomarker was found to be associated with HIV antibody levels, nor were any immune activation biomarkers, even when adjusting for pre-ART biomarker level (**Table 3**), indicating a lack of evidence for HIV antibodies contributing to on-ART inflammation and immune activation under long-term viral suppression.

**Table 3.**
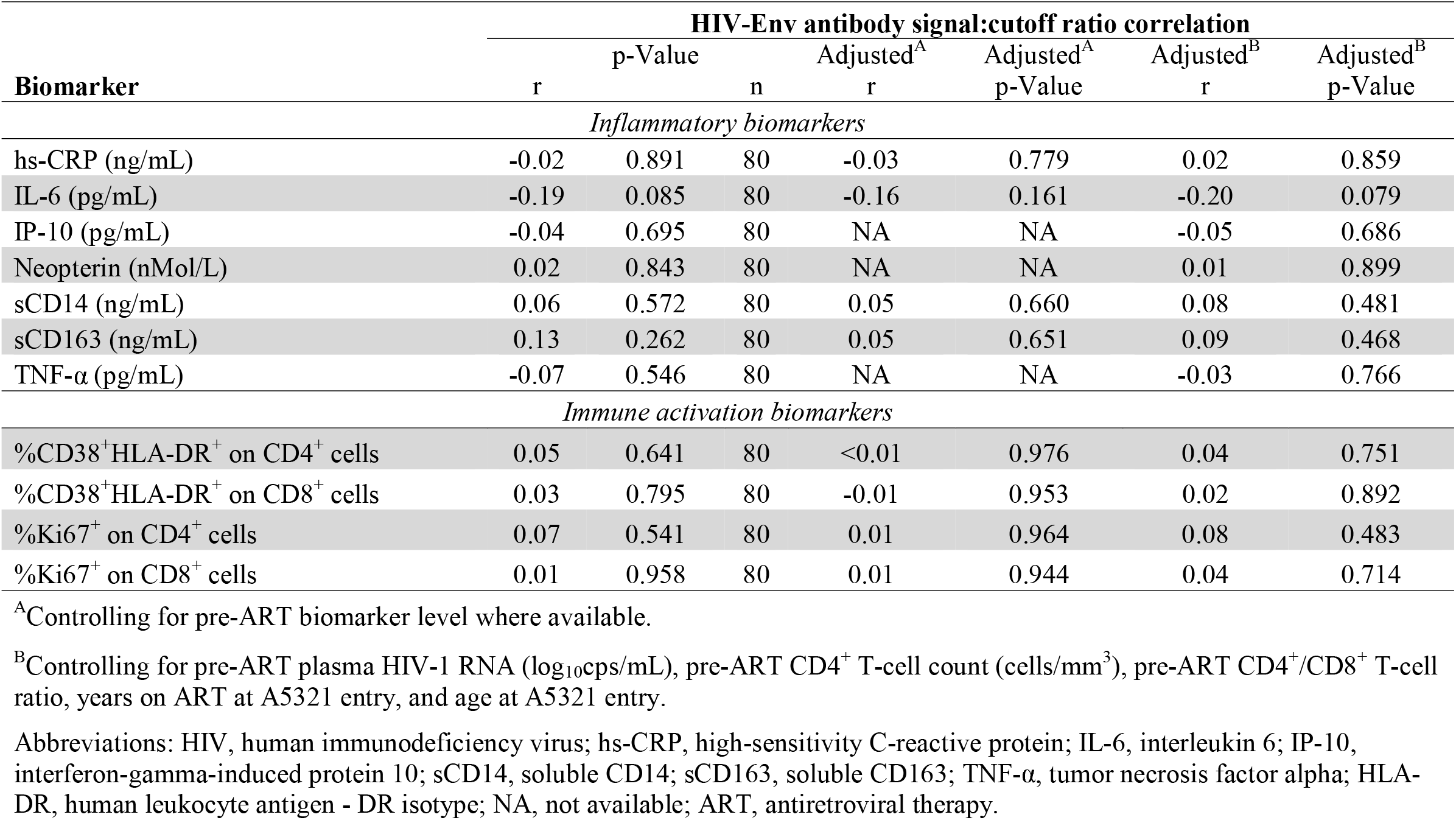
Spearman correlations between HIV-Env-specific antibody levels with inflammatory and immune activation biomarkers at A5321 study entry

### HIV-Specific Immune Responses do not Mediate an Association Between Reservoir Parameters and Levels of Inflammation or Immune Activation

A previous study reported a lack of correlations between measures of HIV persistence, including CA-DNA, CA-RNA, and plasma HIV RNA by iSCA, with levels of inflammatory and immune activation biomarkers at year 4 on therapy in the A5321 cohort [12]. We extended these results to analyses at the last longitudinal time point currently available on the cohort (A5321 study entry), a median of 7 (range 4 to 15) years on ART. CA-DNA levels were not associated with any inflammatory or immune activation biomarker, including when associations were adjusted for pre-ART biomarker level (**Table S2**). Likewise, CA-RNA levels were not associated with any inflammatory or immune activation biomarker (**Table S3**), nor was the CA-RNA:CA-DNA ratio (a surrogate of active replication) (**Table S4**). Interestingly, plasma HIV RNA levels by iSCA were weakly correlated with CD8^+^ T-cell activation as measured by %CD38^+^HLA-DR^+^ CD8^+^ cells (r = 0.25, p = 0.027), which remained significant after controlling for potential confounding by pre-ART plasma viral load, CD4^+^ T-cell count, and CD4^+^/CD8^+^ T-cell ratio, years on ART at A5321 entry, and age at A5321 entry (adjusted r = 0.24, p = 0.037) (**Figure S2 and Table S5**). This association also remained significant, and similar in magnitude, when adjusting for pre-ART %CD38^+^HLA-DR^+^ CD8^+^ cells (adjusted r = 0.26, p = 0.024) (**Table S5**), suggesting that on-ART low-level viremia – undetectable by commercial assays but detectable by single-copy assay – is biologically associated with increased CD8^+^ T-cell activation, regardless of a participant’s baseline level of CD8^+^ T-cell activation prior to ART initiation.

We next sought to determine if the association between plasma HIV RNA by iSCA with CD8^+^ T-cell activation was mediated by ongoing HIV-specific immune responses. In order to test this, we performed statistical mediation analysis using structural equation modeling. In mediation analysis, the objective is to determine if a third variable, Z, represents the causal mechanism (at least in part) through which the independent variable, X, influences the dependent variable, Y – indicated by a significant indirect effect of X through Z on Y [21,22]. SEM is a multivariate technique which also allows for assessing the fit of mediation models – an important indicator of the consistency of the hypothesized mediation model with the actual data – and for comparison of different mediation models using goodness-of-fit statistics [21,23]. We used two goodness-of-fit criteria to evaluate models: the root mean square error of approximation (RMSEA, with a value ≤0.06 supporting the hypothesized model [24]), and the Bentler-Bonnet Normed Fit Index (NFI, with a value ≥0.95 supporting the hypothesized model [24]); additionally, we used the Bayesian information criterion (BIC) to compare and select models, with a ΔBIC >10 taken to indicate a meaningful difference (a lower BIC is better) [25]. We generated models with plasma HIV RNA via iSCA as the independent variable (X), all measured immune activation biomarkers as the outcome variables (Y1-Y4), and either magnitudes of HIV-specific T-cell responses (against each gene product, separately, as well as summed HIV responses) or HIV antibody levels as the mediating variable (M). We present here results of summed HIV T-cell responses and HIV antibody levels as the mediating variables based on BIC statistics. Both SEM models suggested an inconsistency of the hypothesized mediation models with the actual data, with an RMSEA of 0.50 for both SEM models, and NFI values of 0.10 (summed HIV responses) and 0.08 (HIV antibody levels) (**Figure 1**), meaning that the variables are likely not related in the ways we hypothesized. While there was a significant direct effect of plasma HIV RNA via iSCA on CD8^+^ T-cell activation (but not on any other immune activation biomarker, consistent with the correlation results), there was no significant indirect effect through the mediating variable for either summed HIV T-cell responses or HIV antibody levels (**Figure 1 and Table 4**). Taken together, these results indicate that on-ART HIV-specific adaptive immune responses do not mediate or influence the relationship between plasma HIV RNA via iSCA with CD8^+^ T-cell activation.

**Table 4.** Results of mediation analyses for associations between iSCA with CD8^+^ T-cell activation and other immune activation biomarkers

| Independent Variable | Mediating Variable | Outcome Variable | Total Effect | Total Effect p-Value | Direct Effect | Direct Effect p-Value | Indirect Effect | Indirect Effect p-Value |
| --- | --- | --- | --- | --- | --- | --- | --- | --- |
| *Sum HIV-specific T-cell responses* |  |  |  |  |  |  |  |  |
| iSCA | Sum HIV IFN-γ ELISPOT | %CD38<sup>+</sup>HLA-DR<sup>+</sup> CD4<sup>+</sup> cells | 0.086 | 0.446 | 0.085 | 0.454 | 0.001 | 0.953 |
| iSCA | Sum HIV IFN-γ ELISPOT | %CD38<sup>+</sup>HLA-DR<sup>+</sup> CD8<sup>+</sup> cells | **0.244** | **0.022** | **0.231** | **0.031** | 0.013 | 0.476 |
| iSCA | Sum HIV IFN-γ ELISPOT | %Ki67<sup>+</sup> CD4<sup>+</sup> cells | -0.083 | 0.462 | -0.085 | 0.454 | 0.002 | 0.882 |
| iSCA | Sum HIV IFN-γ ELISPOT | %Ki67<sup>+</sup> CD8<sup>+</sup> cells | -0.064 | 0.571 | -0.061 | 0.590 | -0.003 | 0.857 |
| *HIV-Env-specific antibody levels* |  |  |  |  |  |  |  |  |
| iSCA | HIV-Env signal:cutoff ratio | %CD38<sup>+</sup>HLA-DR<sup>+</sup> CD4<sup>+</sup> cells | 0.086 | 0.446 | 0.085 | 0.446 | <0.001 | 0.969 |
| iSCA | HIV-Env signal:cutoff ratio | %CD38<sup>+</sup>HLA-DR<sup>+</sup> CD8<sup>+</sup> cells | **0.244** | **0.022** | **0.243** | **0.022** | <0.001 | 0.969 |
| iSCA | HIV-Env signal:cutoff ratio | %Ki67<sup>+</sup> CD4<sup>+</sup> cells | -0.083 | 0.462 | -0.083 | 0.458 | <0.001 | 0.969 |
| iSCA | HIV-Env signal:cutoff ratio | %Ki67<sup>+</sup> CD8<sup>+</sup> cells | -0.064 | 0.571 | -0.064 | 0.570 | <0.001 | 0.970 |

All variables rank-transformed.
Significant effects at the $\alpha=0.05$ level are bolded.
Abbreviations: iSCA, integrase single copy assay; HIV, human immunodeficiency virus; IFN-γ , interferon-gamma; HLA-DR, human leukocyte antigen - DR isotype.

**Figure 1.**
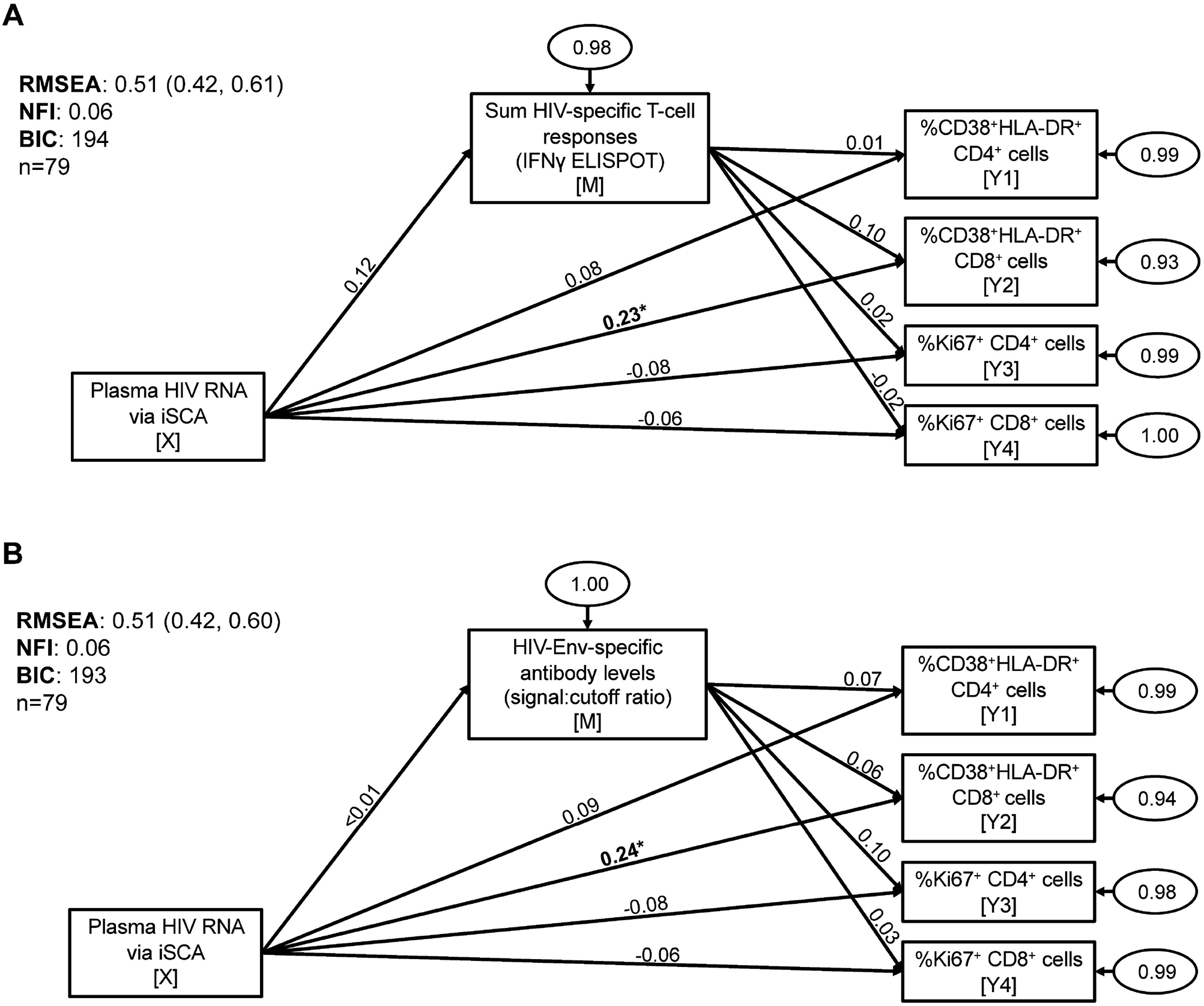
HIV-specific T-cell responses and HIV antibody levels do not mediate the association between plasma HIV RNA levels by iSCA with CD8^+^ T-cell activation. Structural equation modeling was used to test for mediation between plasma HIV RNA via iSCA with markers of T-cell activation (%CD38^+^HLA-DR^+^ CD4^+^ and CD8^+^ cells) and T-cell cycling (%Ki67^+^ CD4^+^ and CD8^+^ cells), mediated by either summed HIV-specific T-cell responses (measured by IFN-γ ELISPOT assay) (*A*) or HIV-Env-specific antibody levels (measured by LS-VITROS signal:cutoff ratio) (*B*). Path model diagrams are depicted, with path coefficients representing standardized effect estimates using rank-transformed data; statistically significant path coefficients are bolded. Circled numbers represent unexplained (error) variances. × indicates the independent variable, M the mediating variable, and Y the outcome variable. Values for goodness-of-fit statistics including RMSEA (with 95% confidence interval), NFI, and BIC are shown. iSCA, integrase single-copy assay; ELISPOT, enzyme-linked immune absorbent spot; RMSEA, root mean square error of approximation; NFI, Bentler-Bonnet Normed Fit Index; BIC, Bayesian information criterion. * p<0.05, ** p<0.01, *** p<0.001.

Next, we tested whether on-ART HIV-specific immune responses mediate an association between HIV reservoir size (by CA-DNA) with levels of inflammation and immune activation. Although CA-DNA levels were not associated with any biomarker in correlation analyses, it is possible for mediating variables to be causally between × and Y, even if X and Y are not directly associated – and X can affect Y indirectly in the absence of a detectable total effect [26]. For example, if the recognition of an HIV-infected cell by an HIV-specific T-cell resulted in the production of IFN-γ, which drove production of IFN-γ-induced protein 10, then the association between infected cells (CA-DNA) and IP-10 would be mediated by HIV-specific T-cells. Such a relationship between CA-DNA and IP-10 might only be revealed if HIV-specific T-cells are considered. We present here results of HIV-Nef/Tat/Rev-specific T-cell responses and HIV antibody levels as the mediating variables based on BIC statistics. When levels of inflammation were considered as outcome variables, both SEM models suggested an inconsistency of the hypothesized mediation models with the actual data, with an RMSEA of 0.17 for both SEM models, and NFI values of 0.15 (Nef/Tat/Rev T-cell responses) and 0.19 (HIV antibody levels) (**Figure S3**). There was a significant effect of CA-DNA levels on both Nef/Tat/Rev responses and HIV antibody levels (**Figure S3**), consistent with the correlations reported in our prior publications [14,15]. There were no significant indirect effects of CA-DNA on inflammatory biomarkers through either Nef/Tat/Rev responses or HIV antibody levels as the mediators (**Figure S3 and Table S6**). Similarly, there were no significant indirect effects of CA-DNA on immune activation biomarkers through either of the mediators tested (**Figure S4 and Table S7**). Taken together, these results indicate that on-ART HIV-specific immune responses do not mediate or influence an association between reservoir size with levels of inflammation and immune activation.

## Discussion

This is the first study to our knowledge assessing the relationships between HIV-specific immune responses with clinically relevant inflammation and immune activation indices – including plasma IL-6 (associated with risk for non-AIDS-defining cancers, cardiovascular disease, renal disease, frailty, and all-cause mortality [27–33]), hsCRP (cardiovascular disease, incident diabetes, mortality [27,29,34,35]), IP-10 (associated with a metric of multimorbidity and mortality [36]), sCD14 (chronic obstructive pulmonary disease, neurocognitive impairment, frailty, mortality [33,37–40]), sCD163 (neurocognitive impairment [41]), and TNF-α (renal disease, frailty [31,33]) – in a cohort of chronic progressors on long-term ART. Our findings indicate that neither HIV-specific T-cell responses, assessed by *ex vivo* IFN-γ production, nor HIV antibody levels influence on-ART levels of inflammation and immune activation. Additionally, we report here that magnitudes of IFN-γ-producing CMV-pp65-specific T-cell responses are not associated with inflammation and immune activation in PWH on long-term ART, consistent with another report assessing CMV-specific T-cell responses by intracellular cytokine staining in n=56 PWH [42]. These latter results add to the literature on CMV, as elevated circulating CD8^+^ T-cell numbers and markers of inflammation and coagulation have been linked to CMV coinfection in PWH [10], but mechanisms are not fully understood. The findings presented here suggest this is not driven by CMV-specific T-cell responses – at least to the pp65 antigen as measured by IFN-γ ELISPOT in the peripheral blood. The association between on-ART plasma HIV levels by iSCA with CD8^+^ T-cell activation differs from the lack of association that we reported previously at an earlier on-ART time point [12,43]. We believe it stands to reason that such an association could become increasingly prevalent with longer durations of ART, during which very low-level viremia and/or viral blips (undetectable by commercial assays) may become increasingly influential drivers of CD8^+^ T-cell activation. It will, however, be important to confirm this observation in future studies on cohorts with similar or longer durations of ART.

Major strengths of our study include that participants had many years of sustained viral suppression prior to entry into the cohort, allowing us to assess the relationships between immune responses with inflammation and immune activation without the potential confounding of appreciable residual viremia or virologic failure. Additionally, longitudinal measures provided for stronger inference by allowing us to adjust for pre-ART values to address the confounding that can occur in cross-sectional studies. The mediation modeling approach also provides a strong inference framework, as it does not rely on a significant total effect between the independent variable and outcome variable to detect meaningful relationships between multiple variables (which may be missed in regression or correlation analyses). Limitations to our study include that we only assessed magnitudes of IFN-γ production but not other qualities of T-cell responses (such as cytotoxicity), which could be important factors related to inflammation and immune activation. Similarly, we only assessed HIV antibody levels but not other functional qualities of antibodies. We also did not measure CMV antibody levels and thus were not able to similarly compare anti-CMV T-cell responses with antibody responses, nor did we measure CMV CA-DNA levels. Immune responses and virologic parameters were only measured in peripheral blood, which may not fully reflect antigen-specific responses in tissues. Additionally, T-cell activation and proliferation measures were performed on total CD4^+^ and CD8^+^ T-cells but not specific cellular subsets, such as HIV-specific or CMV-specific T-cells. Differing measures of the HIV reservoir, such as viral outgrowth or intact vs. defective HIV DNA levels, may yield additional insights into relationships with both anti-HIV immune responses and inflammation.

Delineating the root drivers of persistent inflammation and immune activation in treated HIV infection despite suppressive ART is critical towards the goal of reducing associated non-AIDS-defining morbidities and increased mortality [1]. As there are multiple potential mechanisms by which HIV-specific adaptive immune responses could have contributed to elevation of the clinically-relevant biomarkers we studied, that ongoing responses do not appear to substantially contribute adds evidence to the “die is cast” or “legacy effect” theory proposed by members of the ACTG A5321 study team and others [12,44], which suggests that pathogenic mechanisms that occur before therapy is initiated are the predominate drivers of long-lasting immune dysregulation (i.e. an “immune dysregulation legacy effect”). Mechanisms consistent with this theory include intestinal damage and increased microbial translocation, coinfections, and lymphoid tissue fibrosis [45] – representing important areas for ongoing research into interventional targets to improve the health of PWH.

## Data Availability

All data produced in the present study are available upon formal request to the AIDS Clinical Trials Group through the Proposal Submission System.

## Acknowledgements

The authors would like to thank all members of the ACTG A5321 Team. We also thank Samuel Simmens, Jeanne Jordan, and Manya Magnus for helpful discussions. We gratefully acknowledge the contributions of the study participants, without whom this work would not be possible. The content is solely the responsibility of the authors and does not necessarily represent the official views of the National Institutes of Health.

## References

1. Hunt PW, Lee SA, Siedner MJ. Immunologic Biomarkers, Morbidity, and Mortality in Treated HIV Infection. Journal of Infectious Diseases 2016; 214:S44–S50.

2. Legarth RA, Ahlström MG, Kronborg G, et al. Long-Term Mortality in HIV-Infected Individuals 50 Years or Older. JAIDS Journal of Acquired Immune Deficiency Syndromes 2016; 71:213–218.

3. Wada NI, Bream JH, Martínez-Maza O, et al. Inflammatory Biomarkers and Mortality Risk Among HIV-Suppressed Men: A Multisite Prospective Cohort Study. Clinical Infectious Diseases 2016; 63:984–990.

4. Samji H, Cescon A, Hogg RS, et al. Closing the Gap: Increases in Life Expectancy among Treated HIV-Positive Individuals in the United States and Canada. PLoS ONE 2013; 8:e81355.

5. Zicari S, Sessa L, Cotugno N, et al. Immune Activation, Inflammation, and Non-AIDS Co-Morbidities in HIV-Infected Patients under Long-Term ART. Viruses 2019; 11:200.

6. Brenchley JM, Price DA, Schacker TW, et al. Microbial translocation is a cause of systemic immune activation in chronic HIV infection. Nature Medicine 2006; 12:1365–1371.

7. Marchetti G, Tincati C, Silvestri G. Microbial translocation in the pathogenesis of HIV infection and AIDS. Clinical Microbiology Reviews 2013; 26:2–18.

8. Zevin AS, McKinnon L, Burgener A, Klatt NR. Microbial translocation and microbiome dysbiosis in HIV-associated immune activation. Current Opinion in HIV and AIDS 2016; 11:182–190.

9. Boulougoura A, Sereti I. HIV infection and immune activation. Current Opinion in HIV and AIDS 2016; 11:191–200.

10. Freeman ML, Mudd JC, Shive CL, et al. CD8 T-cell expansion and inflammation linked to CMV coinfection in ART-treated HIV infection. Clinical Infectious Diseases 2016; 62.

11. Martinez-Picado J, Deeks SG. Persistent HIV-1 replication during antiretroviral therapy. Current Opinion in HIV and AIDS 2016; 11:417–423.

12. Gandhi RT, McMahon DK, Bosch RJ, et al. Levels of HIV-1 persistence on antiretroviral therapy are not associated with markers of inflammation or activation. PLOS Pathogens 2017; 13:e1006285.

13. Spudich S, Robertson KR, Bosch RJ, et al. Persistent HIV-infected cells in cerebrospinal fluid are associated with poorer neurocognitive performance. Journal of Clinical Investigation 2019; 129:3339–3346.

14. Thomas AS, Jones KL, Gandhi RT, et al. T-cell responses targeting HIV Nef uniquely correlate with infected cell frequencies after long-term antiretroviral therapy. PLOS Pathogens 2017; 13:e1006629.

15. Keating SM, Jones RB, Lalama CM, et al. Brief Report: HIV Antibodies Decline During Antiretroviral Therapy but Remain Correlated With HIV DNA and HIV-Specific T-Cell Responses. In: Journal of Acquired Immune Deficiency Syndromes. 2019: 594–599.

16. Stevenson EM, Ward AR, Truong R, et al. HIV-specific T cell responses reflect substantive in vivo interactions with antigen despite long-term therapy. JCI Insight 2021; 6.

17. Hong F, Aga E, Cillo AR, et al. Novel Assays for Measurement of Total Cell-Associated HIV-1 DNA and RNA. Journal of Clinical Microbiology 2016; 54:902–911.

18. Cillo AR, Vagratian D, Bedison MA, et al. Improved Single-Copy Assays for Quantification of Persistent HIV-1 Viremia in Patients on Suppressive Antiretroviral Therapy. Journal of Clinical Microbiology 2014; 52:3944–3951.

19. Keating SM, Pilcher CD, Jain V, et al. HIV Antibody Level as a Marker of HIV Persistence and Low-Level Viral Replication. Journal of Infectious Diseases 2017; 216.

20. Conover WJ, Iman RL. Analysis of Covariance Using the Rank Transformation. Biometrics 1982; 38:715.

21. Danner D, Hagemann D, Fiedler K. Mediation analysis with structural equation models: Combining theory, design, and statistics. European Journal of Social Psychology 2015; 45:460–481.

22. Baron RM, Kenny DA. The moderator–mediator variable distinction in social psychological research: Conceptual, strategic, and statistical considerations. Journal of Personality and Social Psychology 1986; 51:1173–1182.

23. Gunzler D, Chen T, Wu P, Zhang H. Introduction to mediation analysis with structural equation modeling. Shanghai archives of psychiatry 2013; 25:390–394.

24. Hu LT, Bentler PM. Cutoff criteria for fit indexes in covariance structure analysis: Conventional criteria versus new alternatives. Structural Equation Modeling 1999; 6.

25. Raftery AE. Bayesian Model Selection in Social Research. Sociological Methodology 1995; 25.

26. Hayes AF. Beyond Baron and Kenny: Statistical mediation analysis in the new millennium. Communication Monographs 2009; 76.

27. Kuller LH, Tracy R, Belloso W, et al. Inflammatory and Coagulation Biomarkers and Mortality in Patients with HIV Infection. PLoS Medicine 2008; 5:e203.

28. Tenorio AR, Zheng Y, Bosch RJ, et al. Soluble Markers of Inflammation and Coagulation but Not T-Cell Activation Predict Non–AIDS-Defining Morbid Events During Suppressive Antiretroviral Treatment. The Journal of Infectious Diseases 2014; 210:1248–1259.

29. Duprez DA, Neuhaus J, Kuller LH, et al. Inflammation, Coagulation and Cardiovascular Disease in HIV-Infected Individuals. PLoS ONE 2012; 7:e44454.

30. Borges ÁH, Silverberg MJ, Wentworth D, et al. Predicting risk of cancer during HIV infection. AIDS 2013; 27:1433–1441.

31. Gupta S, Kitch D, Tierney C, Melbourne K, Ha B, McComsey G. Markers of renal disease and function are associated with systemic inflammation in HIV infection. HIV Medicine 2015; 16:591–598.

32. Borges ÁH, O’Connor JL, Phillips AN, et al. Interleukin 6 Is a Stronger Predictor of Clinical Events Than High-Sensitivity C-Reactive Protein or D-Dimer During HIV Infection. Journal of Infectious Diseases 2016; 214:408–416.

33. Margolick JB, Bream JH, Martínez-Maza O, et al. Frailty and Circulating Markers of Inflammation in HIV+ and HIV™ Men in the Multicenter AIDS Cohort Study. JAIDS Journal of Acquired Immune Deficiency Syndromes 2017; 74:407–417.

34. de Luca A, de Gaetano Donati K, Colafigli M, et al. The association of high-sensitivity c-reactive protein and other biomarkers with cardiovascular disease in patients treated for HIV: a nested case–control study. BMC Infectious Diseases 2013; 13:414.

35. Brown TT, Tassiopoulos K, Bosch RJ, Shikuma C, McComsey GA. Association Between Systemic Inflammation and Incident Diabetes in HIV-Infected Patients After Initiation of Antiretroviral Therapy. Diabetes Care 2010; 33:2244–2249.

36. Premeaux TA, Javandel S, Hosaka KRJ, et al. Associations Between Plasma Immunomodulatory and Inflammatory Mediators With VACS Index Scores Among Older HIV-Infected Adults on Antiretroviral Therapy. Frontiers in Immunology 2020; 11.

37. Justice AC, Freiberg MS, Tracy R, et al. Does an Index Composed of Clinical Data Reflect Effects of Inflammation, Coagulation, and Monocyte Activation on Mortality Among Those Aging With HIVã Clinical Infectious Diseases 2012; 54:984–994.

38. Sandler NG, Wand H, Roque A, et al. Plasma Levels of Soluble CD14 Independently Predict Mortality in HIV Infection. The Journal of Infectious Diseases 2011; 203:780–790.

39. Hunt PW, Sinclair E, Rodriguez B, et al. Gut Epithelial Barrier Dysfunction and Innate Immune Activation Predict Mortality in Treated HIV Infection. The Journal of Infectious Diseases 2014; 210:1228–1238.

40. Attia EF, Akgün KM, Wongtrakool C, et al. Increased Risk of Radiographic Emphysema in HIV Is Associated With Elevated Soluble CD14 and Nadir CD4. Chest 2014; 146:1543–1553.

41. Burdo TH, Weiffenbach A, Woods SP, Letendre S, Ellis RJ, Williams KC. Elevated sCD163 in plasma but not cerebrospinal fluid is a marker of neurocognitive impairment in HIV infection. AIDS 2013; 27:1387–1395.

42. Ballegaard V, Brændstrup P, Pedersen KK, et al. Cytomegalovirus-specific T-cells are associated with immune senescence, but not with systemic inflammation, in people living with HIV. Scientific Reports 2018; 8:3778.

43. Gandhi RT, Zheng L, Bosch RJ, et al. The Effect of Raltegravir Intensification on Low-level Residual Viremia in HIV-Infected Patients on Antiretroviral Therapy: A Randomized Controlled Trial. PLoS Medicine 2010; 7:e1000321.

44. Centlivre M, Sala M, Wain-Hobson S, Berkhout B. In HIV-1 pathogenesis the die is cast during primary infection. AIDS 2007; 21:1–11.

45. Zeng M, Smith AJ, Wietgrefe SW, et al. Cumulative mechanisms of lymphoid tissue fibrosis and T cell depletion in HIV-1 and SIV infections. Journal of Clinical Investigation 2011; 121.

