## Supplementary figures and images for "No Evidence that Ongoing HIV-Specific Immune Responses Contribute to Persistent Inflammation and Immune Activation in Persons on Long-Term ART"

### Supplemental Tables 1-7

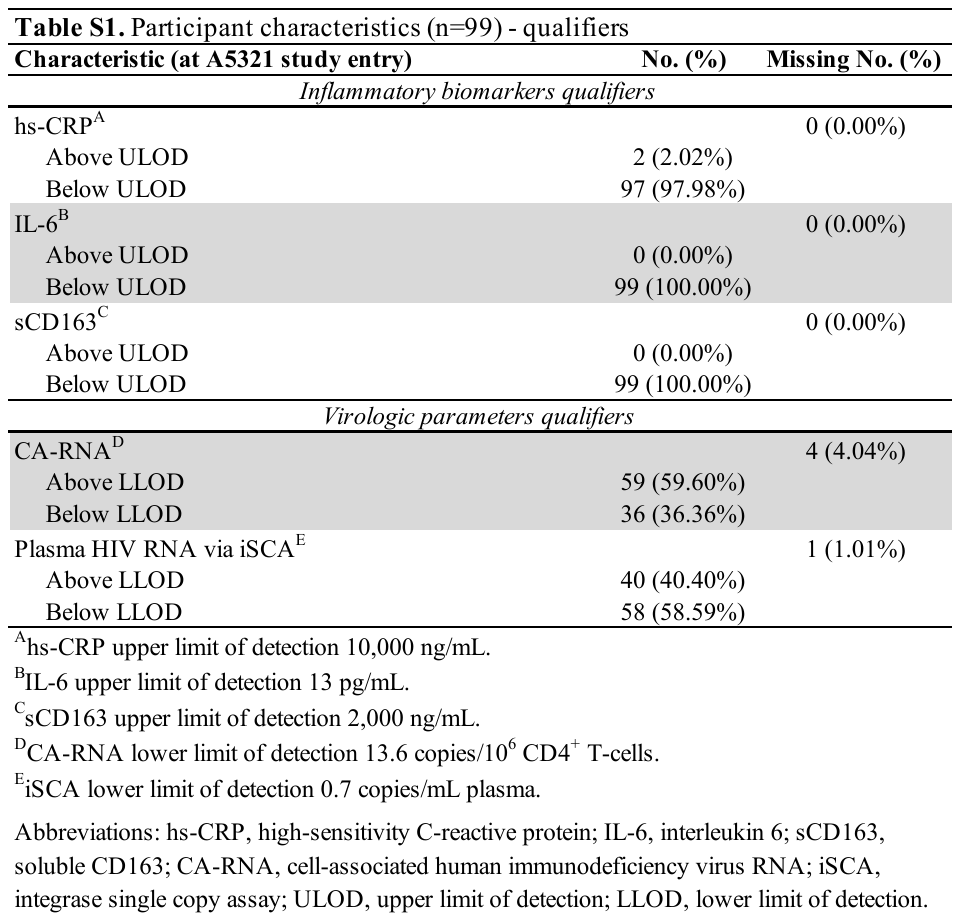

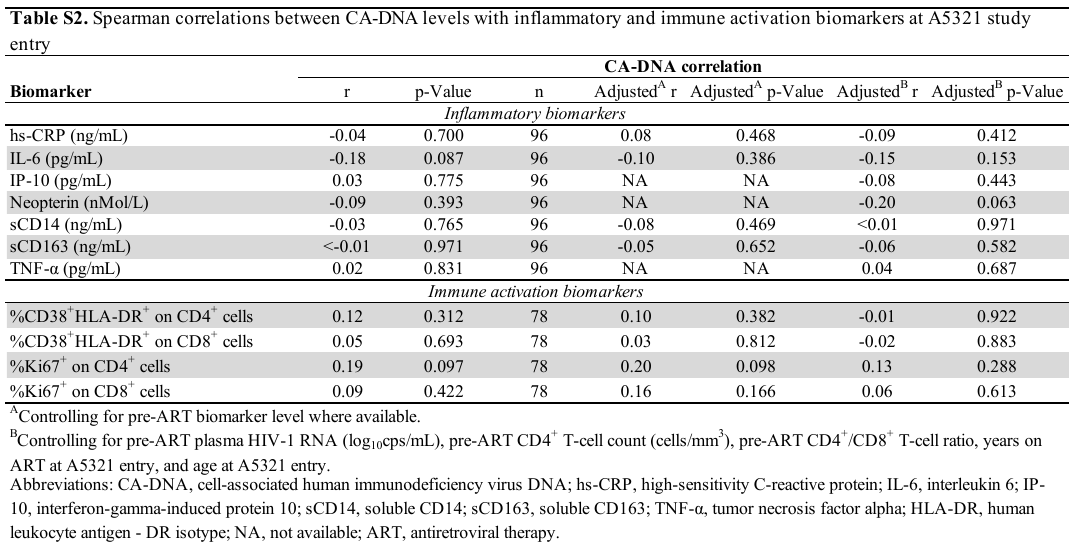

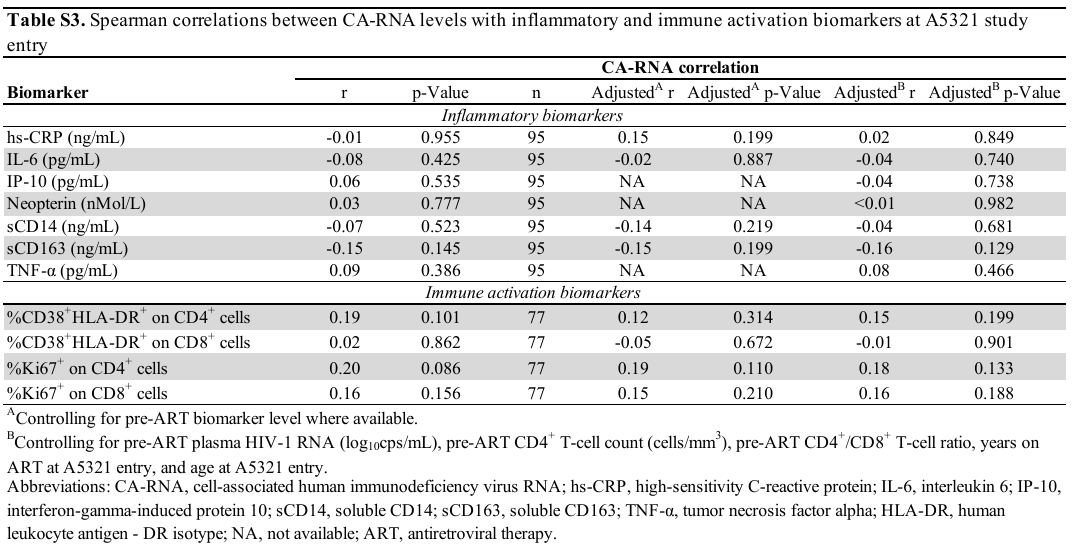

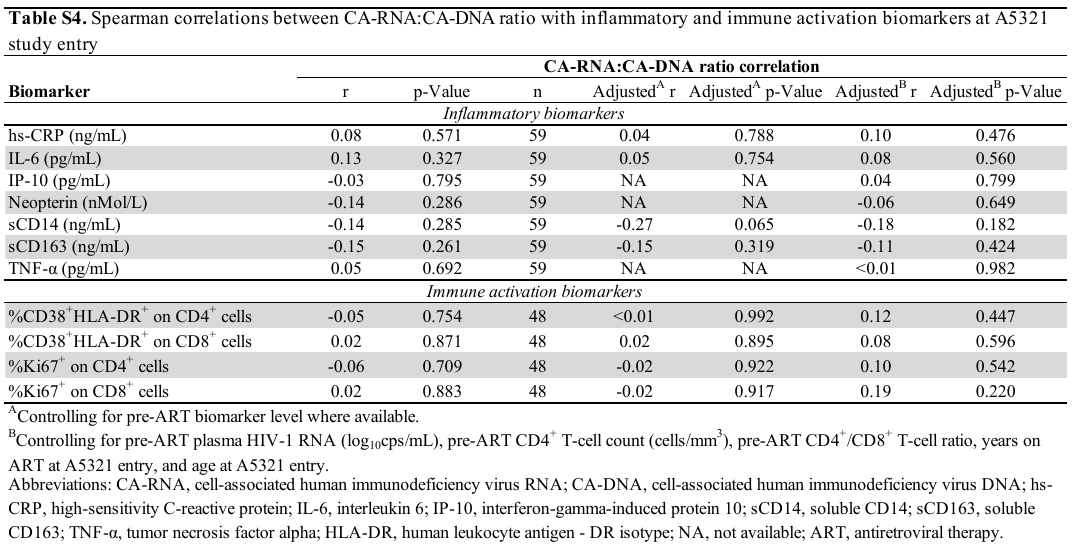

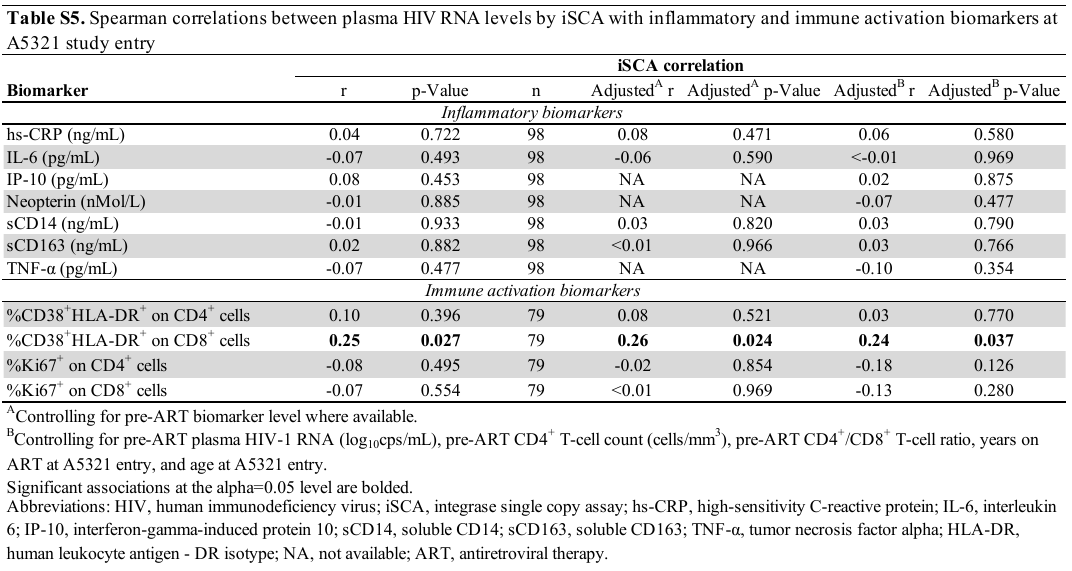

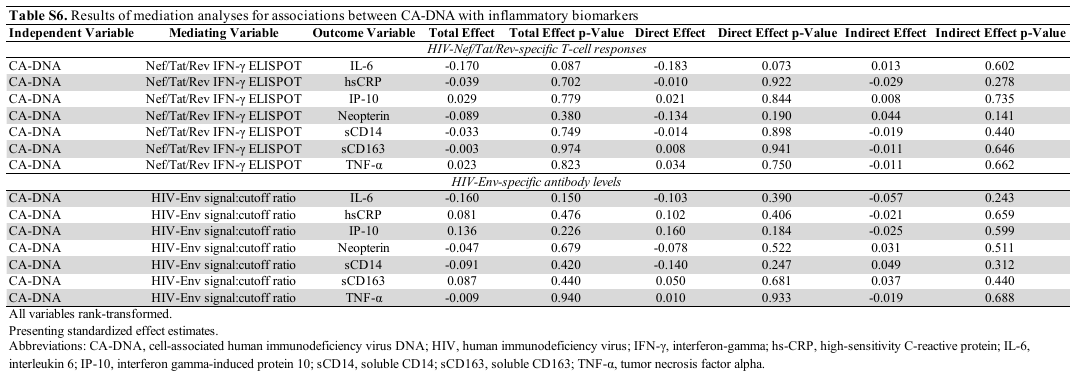

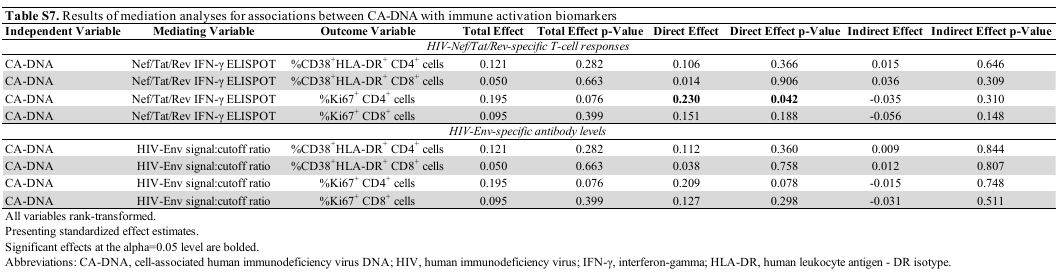
