## Supplemental Figures 1-4 for "No Evidence that Ongoing HIV-Specific Immune Responses Contribute to Persistent Inflammation and Immune Activation in Persons on Long-Term ART"

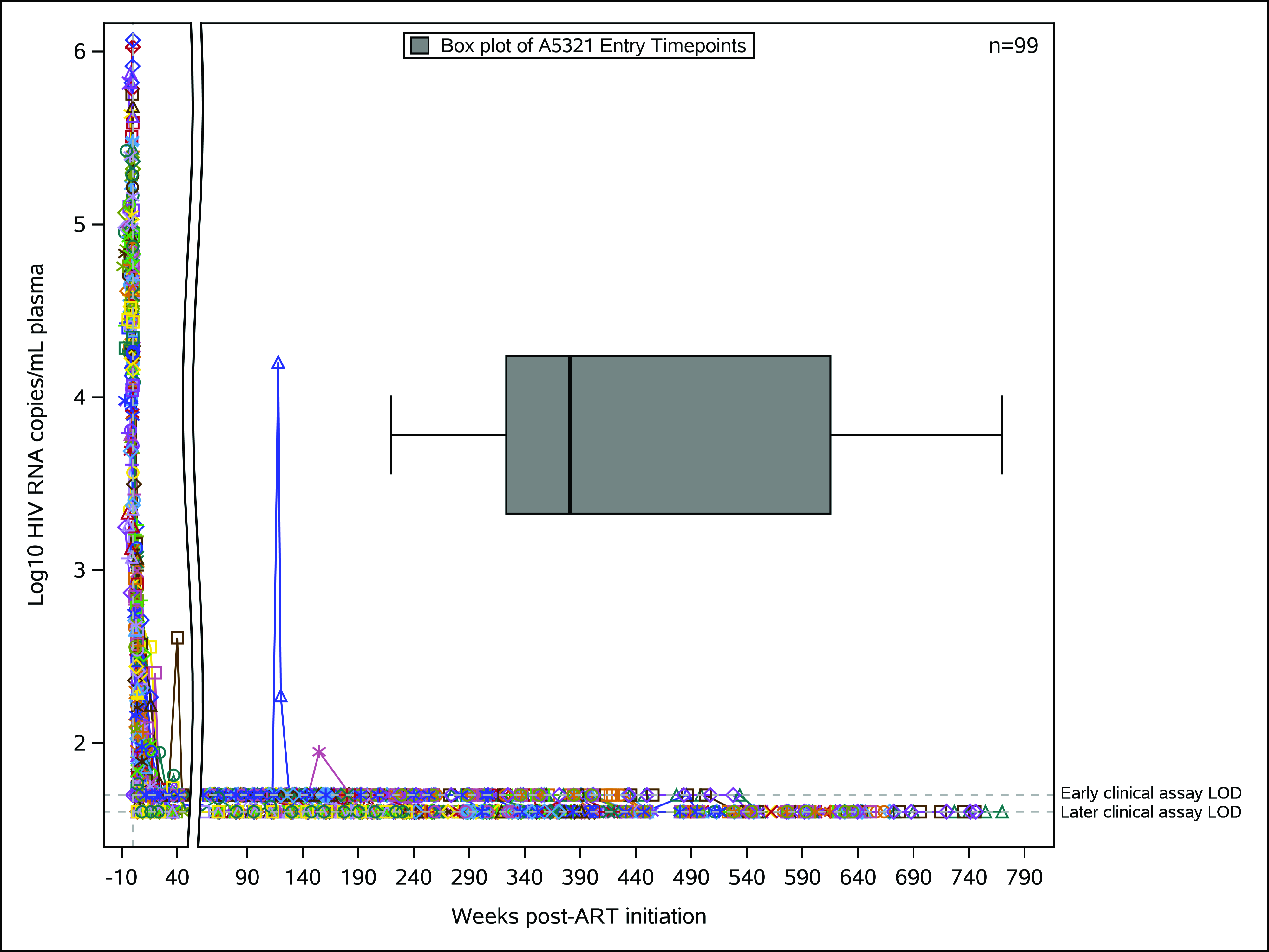

**Figure S1.** ACTG A5321 cohort participants achieved viral suppression by week 48 post-ART initiation and maintained viral suppression prior to A5321 study entry. Graph depicts log_10_ plasma HIV RNA (copies/mL) by clinical commercial assays for ACTG A5321 cohort study participants included in this study (n=99), followed from pre-ART initiation (ART initiated in other ACTG clinical trials) through to A5321 study entry. Lower LOD for early clinical assays was 50 copies/mL, and for later clinical assays 40 copies/mL. Colored lines represent individual participants, with symbols indicating each clinical viral load measurement. X-axis break shows time post-ART initiation when all participants achieved initial viral suppression (48 weeks). Box plot shows the distribution of participants’ A5321 study entry timepoints relative to weeks post-ART initiation (minimum, Q1, median, Q3, maximum). ACTG, AIDS Clinical Trials Group; ART, antiretroviral therapy; LOD, limit of detection.

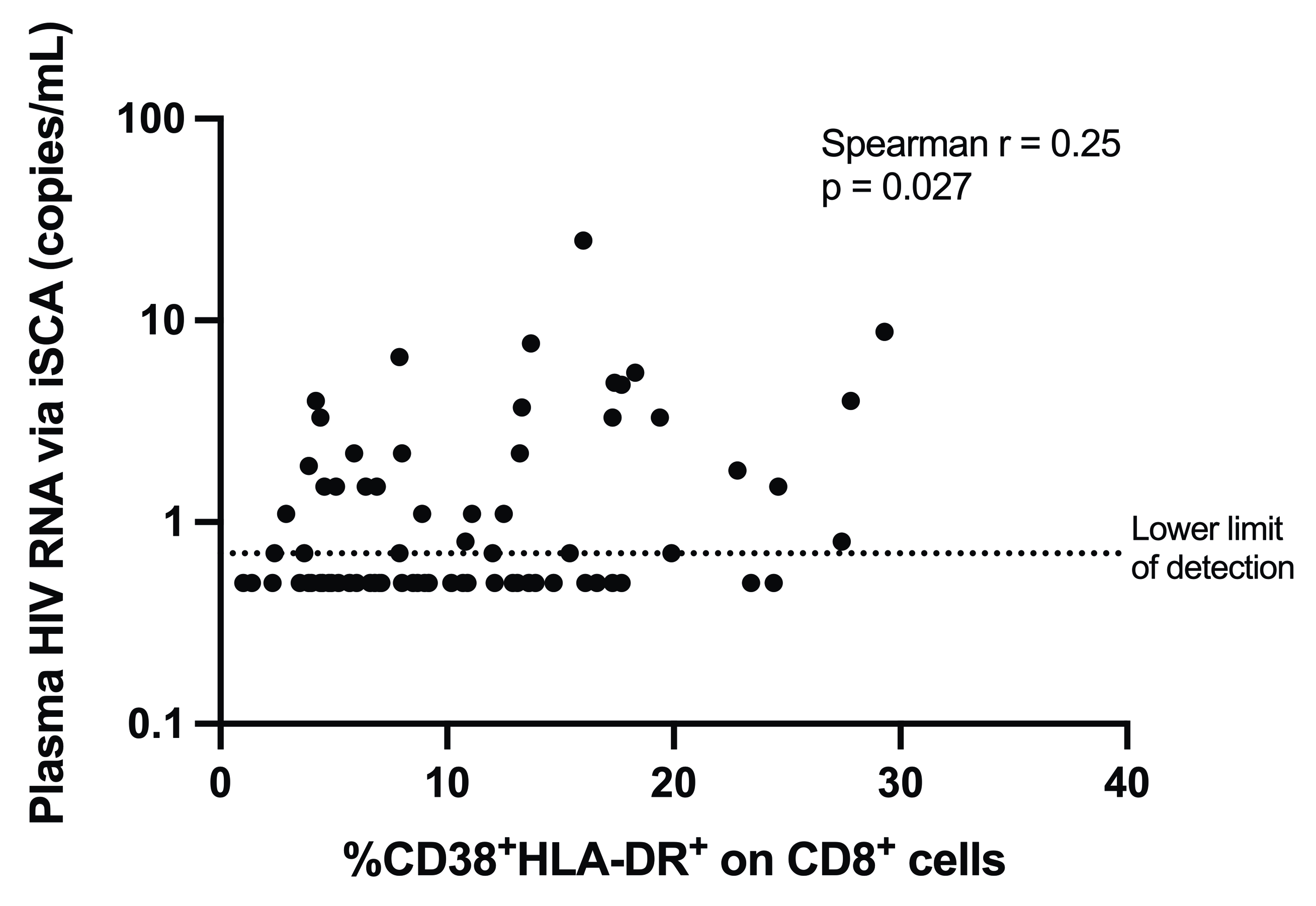

**Figure S2.** Plasma HIV RNA levels by iSCA correlate with CD8^+^ T-cell activation at A5321 study entry. Spearman correlation between plasma HIV RNA via iSCA with CD8^+^ T-cell activation (%CD38^+^HLA-DR^+^ CD8^+^ cells) in A5321 cohort participants (n=79) at study entry. The lower limit of detection for the iSCA assay was 0.7 copies/mL (measured in 5mL plasma). iSCA, integrase single-copy assay.

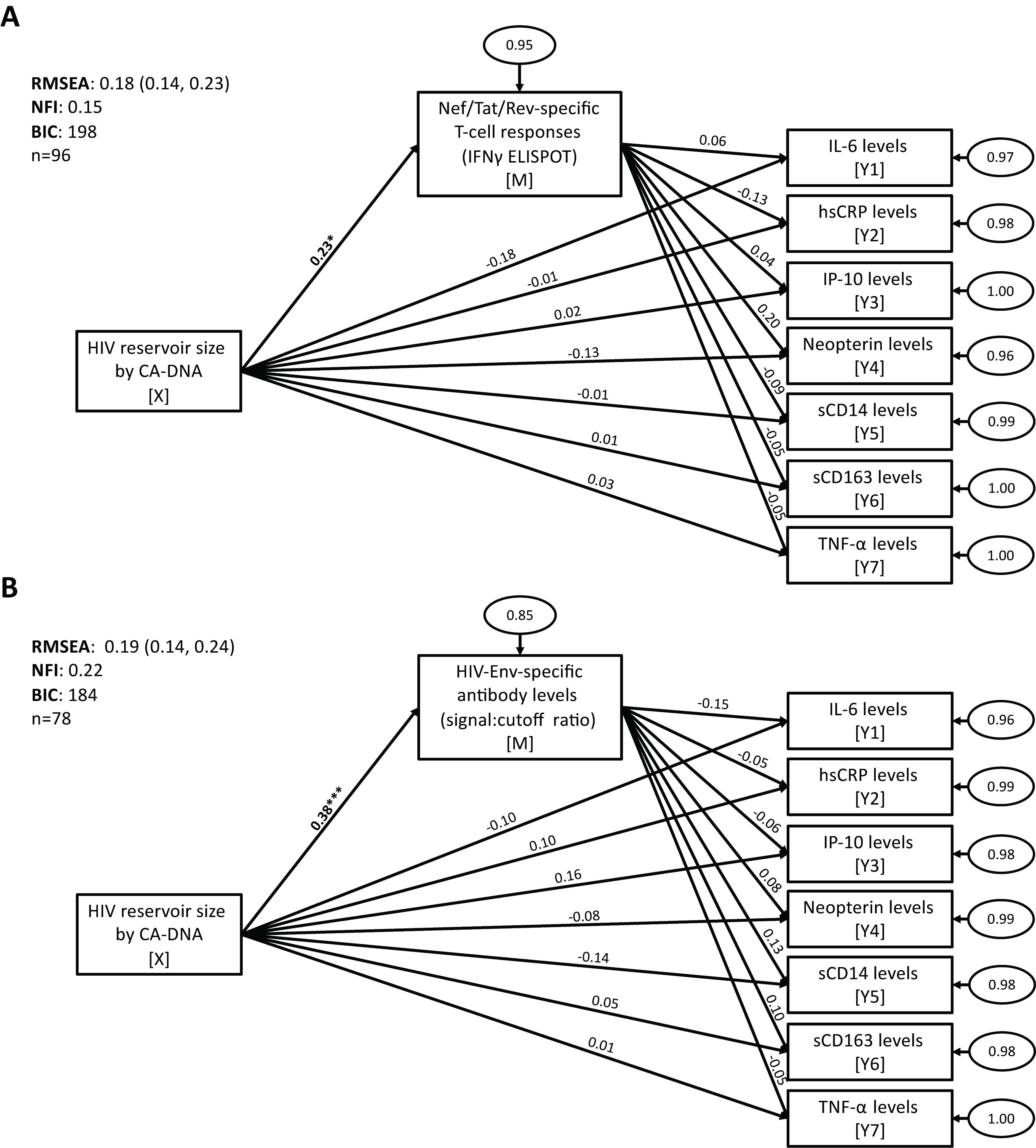

**Figure S3.** HIV-specific T-cell responses and HIV antibody levels do not influence an association between on-ART HIV reservoir size with markers of inflammation. Structural equation modeling was used to test for mediation between CA-DNA levels with markers of inflammation (IL-6, hsCRP, IP-10, neopterin, sCD14 sCD163, and TNF-α), mediated by either HIV-Nef/Tat/Rev-specific T-cell responses (measured by IFN-γ ELISPOT assay) (*A*) or HIV-Env-specific antibody levels (measured by LS-VITROS signal:cutoff ratio) (*B*). Path model diagrams are depicted, with path coefficients representing standardized effect estimates using rank-transformed data; statistically significant path coefficients are bolded. Circled numbers represent unexplained (error) variances. X indicates the independent variable, M the mediating variable, and Y the outcome variable. Values for goodness-of-fit statistics including RMSEA (with 95% confidence interval), NFI, and BIC are shown. CA-DNA, cell-associated HIV DNA; ELISPOT, enzyme-linked immune absorbent spot; RMSEA, root mean square error of approximation; NFI, Bentler-Bonnet Normed Fit Index; BIC, Bayesian information criterion. * p<0.05, ** p<0.01, *** p<0.001.

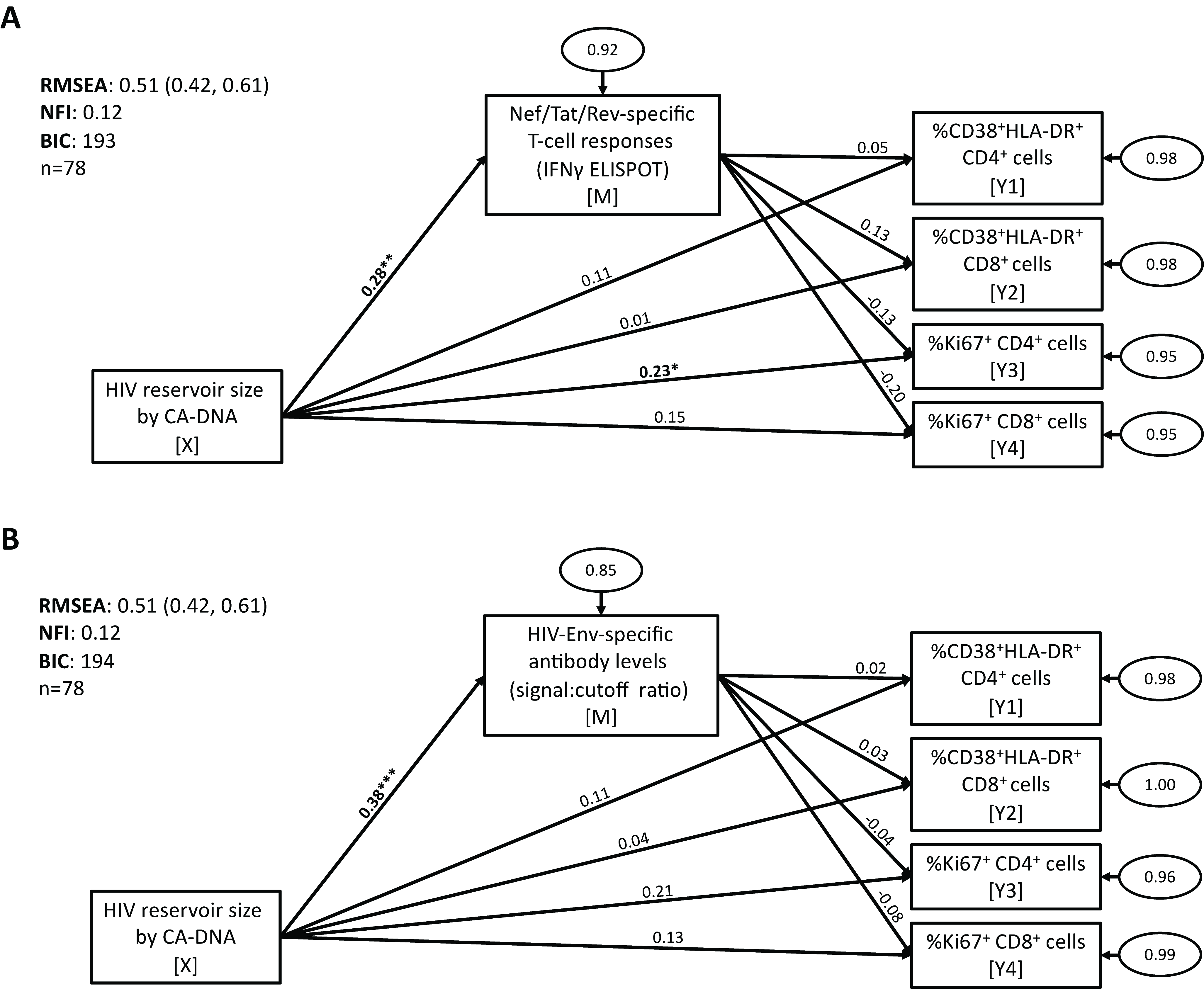

**Figure S4.** HIV-specific T-cell responses and HIV antibody levels do not influence an association between on-ART HIV reservoir size with markers of T-cell activation or cycling. Structural equation modeling was used to test for mediation between CA-DNA levels with markers of T-cell activation (%CD38^+^HLA-DR^+^ CD4^+^ and CD8^+^ cells) and T-cell cycling (%Ki67^+^ CD4^+^ and CD8^+^ cells), mediated by either HIV-Nef/Tat/Rev-specific T-cell responses (measured by IFN-γ ELISPOT assay) (*A*) or HIV-Env-specific antibody levels (measured by LS-VITROS signal:cutoff ratio) (*B*). Path model diagrams are depicted, with path coefficients representing standardized effect estimates using rank-transformed data; statistically significant path coefficients are bolded. Circled numbers represent unexplained (error) variances. X indicates the independent variable, M the mediating variable, and Y the outcome variable. Values for goodness-of-fit statistics including RMSEA (with 95% confidence interval), NFI, and BIC are shown. CA-DNA, cell-associated HIV DNA; ELISPOT, enzyme-linked immune absorbent spot; RMSEA, root mean square error of approximation; NFI, Bentler-Bonnet Normed Fit Index; BIC, Bayesian information criterion. * p<0.05, ** p<0.01, *** p<0.001.
